# Determinants and Patterns of Contraceptive Use among Sexually Active Women living with HIV in Ibadan, Nigeria

**DOI:** 10.1101/2023.09.17.23295687

**Authors:** Folahanmi T. Akinsolu, Zaniab O. Adegbite, Samuel Bankole, Abisola Lawale, Ifeoluwa E. Adewole, Olunike R. Abodunrin, Mobolaji T. Olagunju, Oluwabukola M. Ola, Abel Chukwuemeka, Aisha O. Gambari, Diana W. Njuguna, Jacinta N. Nwogu-Attah, Abideen O. Salako, Akim T. Lukwa, Ebiere Herbertson, George U. Eleje, Oliver C. Ezechi

## Abstract

**Background:** Contraception is a strategy to meet the family planning goals of women living with human immunodeficiency virus (WLHIV) as well as to reduce the transmission of HIV. There is limited data from Nigeria, where HIV prevalent is the second-largest in the world. This study aimed to examine contraceptive use and identify factors influencing its use among sexually active WLHIV in Ibadan, Nigeria.

**Methods:** A facility-based cross-sectional study was conducted involving 443 sexually active WLHIV. The inclusion criteria were WLHIV, aged 18–49 years, who asserted being fecund and sexually active. An adopted questionnaire was used to collect data, and the data was analyzed using the Statistical Package for Social Sciences (SPSS) Windows version 25. Statistical significance was set at p < 0.05.

**Results:** Among sexually active WLHIV (n = 443), 73.1% used contraceptives, with 26.9% having unmet needs. The results revealed a significant association between employment status and the use of contraceptives (AOR] = 2.150; 95% CI 1.279–3.612 p=0.004); accessibility to contraceptive methods and the use of contraceptives (AOR = 21.483; 95% CI 7.279–63.402 p=0.00). Also, a significant association was found between payment for service and contraceptive use (AOR = 14.343; 95% CI 2.705-76.051; p = 0.003). Previous reactions towards contraceptive use were also significantly associated with contraceptive use (AOR = 14.343; 95% CI 2.705-76.051 p = 0.003). The dual contraceptives usage rate was 30.7%.

**Conclusions:** Although contraceptive use among sexually active WLHIV was high, the study highlighted the need for increased adoption of dual contraceptive methods to mitigate the risk of unintended pregnancy and HIV re-infection among this population. It emphasized the importance of continuous sensitization and counseling services healthcare providers provide to promote contraceptive use among WLHIV.

## Introduction

Globally, there are about 37.7 million Human Immunodeficiency Virus (HIV)-positive individuals worldwide, of which 20.2 million are women living with HIV^1^. It has been determined that Sub-Saharan Africa is the area most severely affected by the HIV epidemic, accounting for more than two-thirds of all HIV infections worldwide. Nigeria has the second-largest HIV epidemic in the world and the region’s highest incidence of new infections. In Nigeria, 1.7 million people lived with HIV (PLHIV) in 2020^2^. Women can make independent choices about having children and having sex, regardless of their HIV status. Women living with HIV(WLHIV) must be informed, given the freedom to choose a safe, effective method of contraception, and provided with it at nearby health centers. Because it prevents unplanned pregnancies, contraception may also be essential for preventing Mother-to-Child Transmission of HIV^3^. Strengthening contraceptive programs is therefore important to lower the high incidence of unintended pregnancies, which could contribute to the elimination of HIV/AIDS outbreaks by 2030 (the Sustainable Development Goal 3.3)^4^.

There appears to be a substantial unmet need for contraception among WLHIV and contraceptive failure due to the high incidence of unwanted pregnancy and abortion^5^. Nigeria accounts for 70% of sexually active WLHIV, 5.5 million WLHIV births annually, and 15% of the world’s low contraceptive uptake^6^. Furthermore, prior research found that Nigerian WLHIV had a high level of awareness about contraception, but that knowledge did not match their use of contraceptives, which was linked to a high proportion of unwanted pregnancies^7^.

Women who are fecund, sexually active, and who report not wanting any more children or wishing to delay the next child are considered to have unmet needs, according to the WHO^8^. Because ART has increased the overall survival rate of PLHIV, women living with HIV must have defined reproductive life plans that include accessible access to contraception. Most of the research that has been done on contraceptive use among WLHIV has focused on married women^9^. Because of this, sexual activity among sexually active unmarried women puts them at a higher risk of having an unexpected child because of the idea that sex can only take place within the setting of a married relationship. Therefore, there is a lack of reproductive plans for these women through health services^10^. Although the WHO has recommended that WLHIV have the right to choose any method of contraception, similar to HIV-negative women, the choice of contraception in the presence of HIV appears to be more complicated because WLHIV is required to strike a balance between the prevention of unintended pregnancy and HIV transmission^11^. Sexually active WLHIV can also plan their reproductive lives to avoid unwanted pregnancy and enjoy parenthood like their counterparts who are not living with HIV. Hence, this study aims to identify key predictors of contraceptive use among sexually active WLHIV in Ibadan, Oyo State, Nigeria.

## Materials and Methods

### Study Design and Setting

The study adopted a facility-based cross-sectional survey conducted among sexually active WLHIV. The study was conducted in three (3) HIV treatment centers in Oyo State, Nigeria: Adeoyo Maternity Hospital Yemetu, State Hospital Adeoyo Ring Road, and St. Annes Anglican Hospital Molete. This study was carried out between September and November 2022.

### Study population and sampling

A purposive sampling method was adopted to select the health facilities because ART treatment is unavailable in all facilities. The participants for the study were randomly selected from each of the health facilities. Inclusion criteria for this study were WLHIV, who reported to (a) be aged between 18 and 49 years and (b) have been sexually active within the last six months. In this study, a flowchart illustrated that 750 WLHIV were interviewed, of which 443 sexually active WLHIV were analyzed. (See Figure 1)

**Figure 1:**
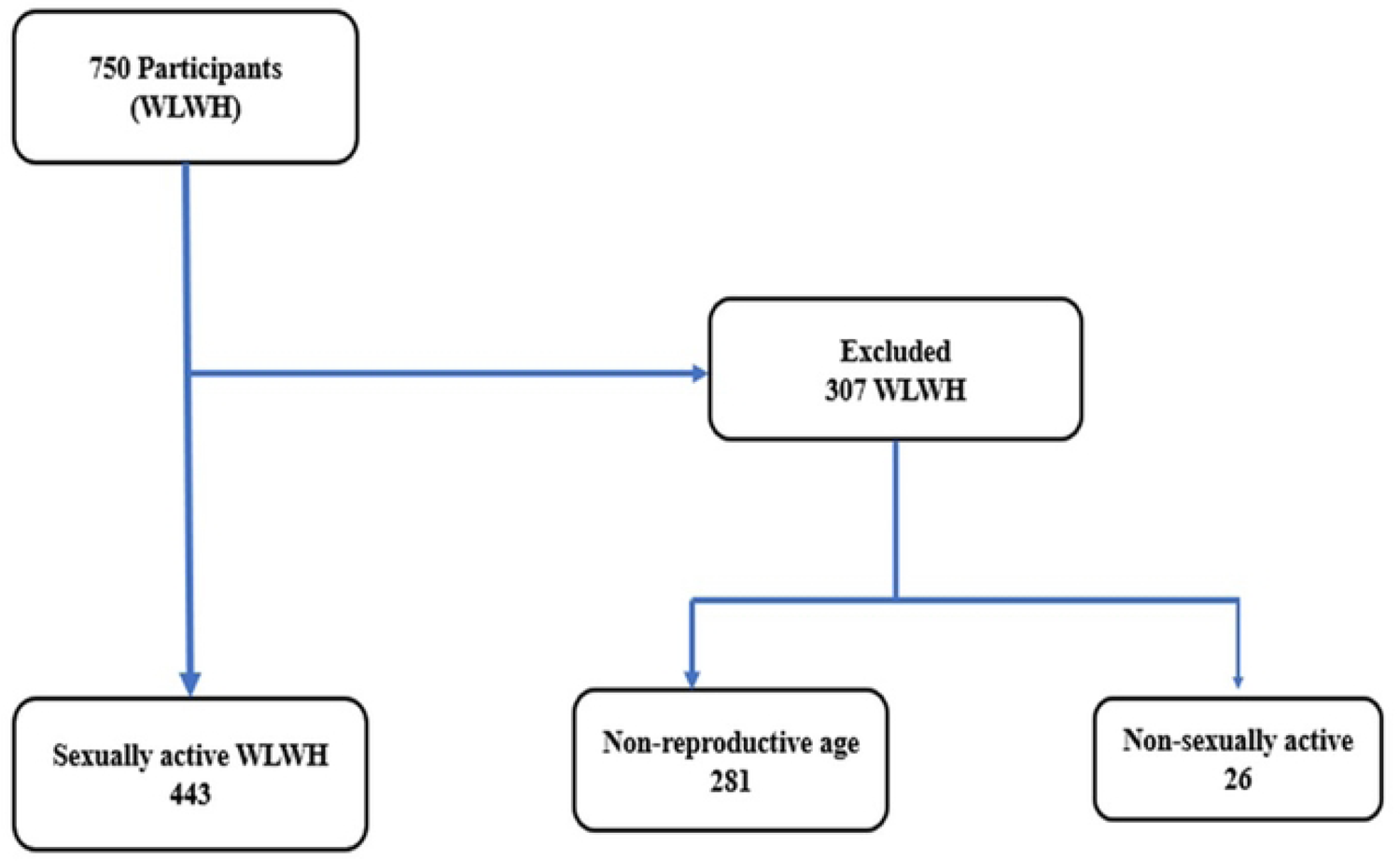
Flow Chart Illustrating the Eligibility Process to Obtain The Final Sample For Analysis of Sexually Active WLHIV

### Data Collection

Data was collected using self-structured and adapted questionnaires from a previous study on contraceptive use among sexually active WLHIV^12^. The questionnaire was divided into three sections: Socio-Demography Data Questionnaire among sexually active WLHIV, Contraceptive Use Questionnaire among sexually active WLHIV, and Perceived factors influencing Contraceptive Use Questionnaire among sexually active WLHIV. The selected participants were those who voluntarily consented to participate. The Research Assistants translated the questionnaires into the language participants understood, Yoruba, for accessible communication.

### Statistical Analysis

The information obtained from the completed questionnaire was processed and analyzed using Statistical Package for Social Science version 25. Descriptive statistics of frequency and proportion were used for the use of contraceptives. Meanwhile, logistic regression was also used to determine the association between various socio-demographic factors and contraceptive use. A P-value less than 0.05 was considered to be statistically significant.

### Ethical Considerations

Ethical approval for this study was obtained from the University Research Ethics Committee (HREC) of Lead City University, Ibadan, Ibadan, Oyo State (LUC-REC/22/124), and Ibadan, Oyo State Ministry of Health Department of Planning Research & Statistics Division (AD 13/479/ 44542A). Official permission was obtained from hospitals included in this study. The consent procedure was conducted in a separate and private room, administered by trained data collectors. Participants were assured that their involvement was entirely voluntary, and they retained the right to decline participation or revoke their consent at any juncture. Significantly, it was stressed that their participation would not impact the medical care they received. Participants were also apprised that the survey might entail sensitive or personal inquiries related to reproductive health concerns, which could be uncomfortable or distressing. Also, participants were explicitly informed that they were under no obligation to answer any question that they found uncomfortable and had the liberty to withdraw from the study or choose not to respond to specific questions at any point. In cases where participants required emotional support, female nurses were available to offer psychological assistance. All collected data were transformed into an anonymized format and stored on laptops protected by passwords throughout the data collection. Furthermore, the data was stored on secure, password-protected computers to ensure confidentiality and security.

## RESULTS

### Socio-demographic Characteristics of Participants

Among 443 sexually active WLHIV eligible for this study, most of the participants, 324 (73.1%), use contraceptives, with a total unmet need of 119 (26.9%). The mean age was 36.88 years (SD= 6.85 years). Most participants (389(87.8%) were married, and 380 (85.8%) identified as Yoruba. From an academic perspective, the majority were secondary school leavers, 199 (44.9%), while 357 (80.6%) of participants were employed, and 214 (48.3 %) of the women reported monthly income of less than 44.71 USD (NGN 33,000). (See Table 1)

**Table 1.** Socio-demographic Characteristics of Study Participants (n =433)

|  | **Contraceptive User**<br>**N=324**<br>**[73.1%]** | **Non-users of**<br>**Contraceptives**<br>**N=119**<br>**[26.9%]** |
| --- | --- | --- |
| **Variable** | **N [%]** | **N [%]** |
| **Age** |  |  |
| Mean ± SD | 36.62±6.70 | 37.59 ± 7.20 |
| **Marital Status** |  |  |
| Married | 290 [74.6] | 99 [25.4] |
| Divorce | 7 [70.0] | 3 [30.0] |
| Widowed | 9 [56.2] | 7 [43.8] |
| Separated | 8 [66.7] | 4 [33.3] |
| Single | 10 [62.5] | 6 [37.5] |
| **Ethnic Group** |  |  |
| **Yoruba** | 285 [75.0] | 95 [25.0] |
| Igbo | 25 [62.5] | 15 [37.5] |
| Hausa | 6 [50.0] | 6 [50.0] |
| Others | 8 [72.7] | 3 [27.3] |
| **Religion** |  |  |
| Christianity | 188 [72.0] | 73 [28.0] |
| Islam | 136 [74.7] | 46 [25.3] |
| **Educational Level** |  |  |
| Primary level | 93 [78.8] | 25 [21.2] |
| Secondary level | 135 [67.8] | 64 [32.2] |
| Tertiary level | 65 [78.3] | 18 [21.7] |
| None | 31 [72.1] | 12 [27.9] |
| **Employment Status** |  |  |
| Employed | 273 [76.5] | 84 [23.5] |
| Unemployed | 51 [59.3] | 35 [40.7] |
| **Type of Partner** |  |  |
| Spouse | 269 [74.9] | 90 [25.1] |
| Steady | 32 [80.0] | 8 [20.0] |
| Causal | 14 [53.8] | 12 [46.2] |
| None | 9 [50.0] | 9 [50.0] |
| **Monthly Income** |  |  |
| < 44.71 USD (NGN 33,000) | 225(77.6) | 65(22.4) |
| ≥ 44.71 USD (NGN 33,000) | 99(64.7) | 54(35.3) |
| **Know Partner's Status** |  |  |
| Yes | 276 [76.9] | 83 [23.1] |
| No | 48 [57.1] | 36 [42.9] |
| **If Yes, Partner Status** |  |  |
| Positive | 111 [77.1] | 33 [22.9] |
| Negative | 161 [78.2] | 45 [21.8] |
| Not Applicable | 52 [55.9] | 41 [44.1] |

### Contraceptive Use among Sexually Active WLHIV

Among sexually active WLHIV, 73.1% used contraceptives. Most participants used male condoms (51.4%), followed by pills (12.50%), and surprisingly, 11.5 % of the sexually active WLHIV used female condoms. (See Figure 2) Additionally, the number of single contraception users was 188 (42.4%), while the number of dual contraception users was 136 (30.7%). (See Table 2)

**Figure 2:**
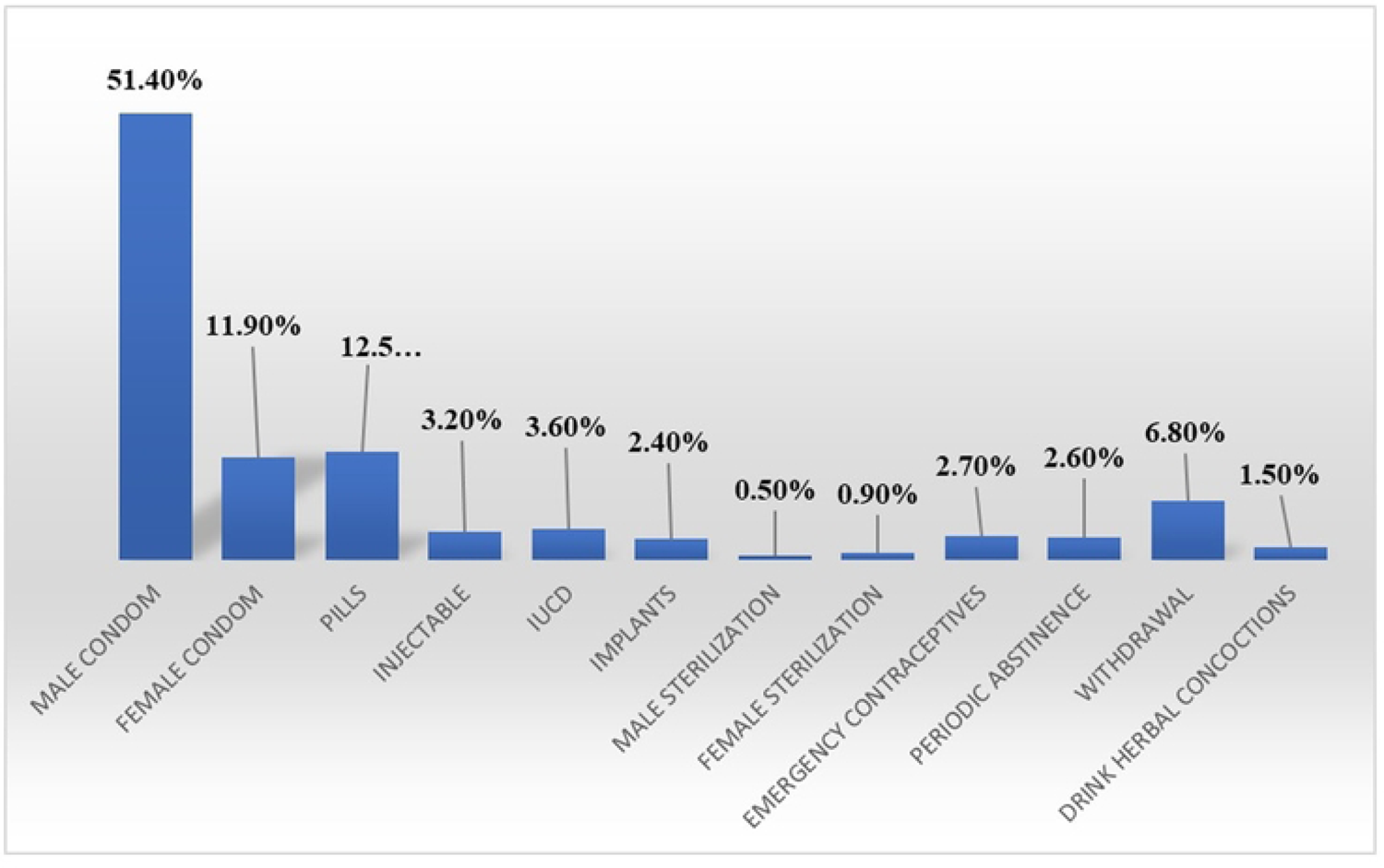
Contraceptive Methods Used Among Sexually Active WLHIV

**Table 2:** Level of Dual Contraceptive Utilization.

| Variable | N [%] |
| --- | --- |
| Dual Contraceptives Usage | 136 [30.7] |
| Single Contraceptive Usage | 188 [42.4] |
| No Usage | 119 [26.9] |

### Factors Associated with Contraceptive Use among Sexually Active WLHIV

After adjusting for the seven significant variables in the logistic regression analysis, only four risk factors had a statistically significant association with contraceptive use among sexually active women living with HIV. These factors are *Employment Status* - unemployed participants were twice times less likely to use contraceptives than their employed counterparts (OR = 2.150, 95% CI 1.279-3.612); *Accessibility to Contraceptive Methods* - participants who obtained their contraceptives outside the ART center were less likely to use contraceptives compared to those who acquired them at the ART center (OR = 21.483, 95% CI 7.279–63.401); *Payment for Service* - participants who had to pay for the service were less likely to use contraceptives than those who did not (OR = 14.343; 95% CI 2.705-76.051); *Previous Negative Reaction to Contraceptives* were less likely to make use of contraceptives (OR = 3.866; 95% CI 0.00). (See Table 4)

**Table 4:** Factors Affecting the Use of Contraceptives among Sexually Active Women Living with HIV.

| Variables | Adjusted Odd Ratio (95% CI) | P value |
| --- | --- | --- |
| **Religion** |  |  |
| Christianity | 0.461(0.176, 1.213) | 0.117 |
| Islam | Ref | Ref |
| **Education Level** |  |  |
| Primary Level | 0.518 (0.162, 1.651) | 0.266 |
| Secondary Level | 1.405(0.438, 4.511) | 0.380 |
| Tertiary Level | 0.354(0.086,1.448) | 0.058 |
| None | Ref |  |
| **Marital status** |  |  |
| Married | 0.661(0.162,1.651) | 0.266 |
| Divorce | 0.490(0.100,2.410) | 0.380 |
| Widowed | 0.184(0.032,1.062) | 0.585 |
| Separated | 1.799(0.184,17.619) | 0.614 |
| Single | Ref |  |
| **Employment Status** |  |  |
| Unemployed | 2.150(1.279,3.612) | 0.004* |
| Employed | Ref |  |
| **Type of Partner** |  |  |
| Spouse | 0.498(0.124,1.993) | 0.233 |
| Steady | 0.102(0.022,0.485) | 0.067 |
| Causal | 0.338(0.035,3.300) | 0.062 |
| None | Ref |  |
| **Get Contraceptives outside ART Center** |  |  |
| Yes | 21.483(7.279,63.402) | 0.00* |
| No | Ref |  |
| **Distance to the health facilities** |  |  |
| ½ to 1 kilometer | 2.669(0.949,0.949) | 0.639 |
| 2 to 3 kilometres | 4.021(1.343,12.036) | 0.304 |
| 4 to 5 kilometres | Ref |  |
| **Payment for Services** |  |  |
| Yes | 14.343(2.705,76.051) | 0.020* |
| No | Ref |  |
| **Adverse Reaction while using Contraceptive** |  |  |
| Yes | Ref |  |
| No | 0.006(0.002,0.20) | 0.000* |

## DISCUSSION

The study underscored the significant use of contraceptives among sexually active women living with HIV (WLHIV) in Ibadan, Nigeria, mirroring the contraceptive adoption rates seen in Ethiopia, where 75% of sexually active WLHIV used contraceptives while the remaining 25% faced unmet contraceptive requirements^12^. This utilization of contraceptives plays a fundamental role in advancing family planning goals and reinforcing the Prevention of Mother-To-Child Transmission (PMTCT) program^13^. Yet, the contrasting findings from Oyo State, Nigeria, remind us that awareness does not always lead to action. Even with high contraceptive knowledge levels among WLHIV, usage rates remained disappointingly low^7^. Male condoms were most popular, trailed by pills and female condoms.

Conversely, less conventional methods like male and female sterilization and herbal mixtures were minimally favored. Compared to data from Togo, these rates were marginally lower^14^. Although dual contraceptive methods could effectively prevent unwanted pregnancies and STIs, their adoption was overshadowed by the preference for single methods^15^.

Several factors are pivotal in determining contraceptive choices^16,17,18^. Employment status stood out, corroborated by Banten Province, Indonesia’s findings, which identified employment as a critical influencer^19^. Similarly, the absence of side effects encouraged continuity in contraceptive use, emphasizing the value of a smooth experience. Contrastingly, the necessity of payments acted as a deterrent, with a study from Uganda linking payment barriers to reduced contraceptive use^17^. Nevertheless, the challenges faced by WLHIV are multi-layered. Sociocultural dynamics heavily influence contraceptive choices^20^. In many African settings, reproductive choices are often determined more by a woman’s partner than by herself. Moreover, the stigma attached to HIV often discourages WLHIV from availing contraceptive services, particularly if healthcare professionals hold biased views^20,21,22,23^.

The types of available contraceptives also matter. The dominant use of male condoms, while indicative of their dual protective nature against STIs and pregnancies, also shows a limitation in contraceptive choices. There’s an evident need for long-acting methods like IUDs and implants, offering women greater autonomy, but their accessibility is often constrained by availability or cost. The role of healthcare infrastructure is undeniable. The inconsistency in contraceptive stock can discourage WLHIV from relying on specific methods, prompting them to settle for less preferred options. Integrating HIV care with contraceptive services might present a more consistent and comprehensive solution^7^.

Comprehensive counseling provides holistic information about contraceptive options, side effects, and effectiveness and can significantly guide WLHIV in making informed choices^24,25^. This approach can demystify misconceptions and align contraceptive choices with individual reproductive and health goals. Lastly, policy frameworks are pivotal. For effective contraceptive adoption among WLHIV, governments and health bodies must craft policies prioritizing their unique challenges, encompassing contraceptive procurement, training of healthcare professionals, and robust monitoring mechanisms.

In essence, while global data paints an overarching picture, addressing the contraceptive needs of WLHIV requires a deeper understanding of the intricate blend of personal, sociocultural, and structural factors. Only a holistic approach, cognizant of these intricacies, can genuinely champion the reproductive rights of every woman.

Self-reported data were used, which may be affected by social desirability and recall bias. The study only focused on women living with HIV and did not include men living with HIV, which limits the understanding of the contraceptive needs of male partners of women living with HIV. The study did not explore the impact of cultural and religious beliefs on contraceptive use, which could significantly influence contraceptive use among this population.

The study’s strengths include a large representative sample of the three facilities and participants, including WLHIV, which was possible due to the nature of the facilities that participated in the study.

## CONCLUSION

This study provides evidence that show high levels of contraceptive use among sexually active WLHIV. However, the study identified the need for greater uptake of dual contraceptive methods to reduce the risk of unwanted pregnancy and HIV re-infection among WLHIV. The study also highlighted various factors, such as employment status, access to contraceptive methods, payment for service, and previous experience with contraceptive use, that influence the use of contraceptives among this population. These findings have important implications for policymakers and healthcare providers seeking to improve reproductive health outcomes and reduce the burden of HIV among WL HIV in Nigeria.

## Funding

“This research received no external funding.”

### Institutional Review Board Statement

The study was conducted following the Declaration of Helsinki and approved by the Ethics Committee of Lead City University, Ibadan (Protocol code: LUC-REC/22/124 and date of approval: September 05, 2022) as well as from the Oyo State Ministry of Health Research Ethics Committee (Protocol code: AD 13/479/ 44542^A^ and date of approval: August 15, 2022).

### Informed Consent Statement

“Informed consent was obtained from all subjects involved in the study.”

## Data Availability

All relevant data are within the manuscript and its Supporting Information files.

## Acknowledgments

This work was supported by grants from Fogarty International Center (FIC) and the National Institute of Health (Funding provided by Fogarty Training Grant: D43TW010934-03). The content is solely the author’s responsibility and does not necessarily represent the official views of the National Institutes of Health.

